# Comprehensive genomic, immunological and clinical analysis of COVID-19 vaccine breakthrough infections: a prospective, comparative cohort study

**DOI:** 10.1101/2021.11.17.21266499

**Authors:** Di Tian, Yang Song, Man Zhang, Yang Pan, Ziruo Ge, Yao Zhang, Xingxiang Ren, Jing Wen, Yanli Xu, Hong Guo, Peng Yang, Zhihai Chen, Wenbo Xu

## Abstract

**Objectives:** As the COVID-19 pandemic is still ongoing and SARS-CoV-2 variants are circulating worldwide, an increasing number of breakthrough infections have been detected despite the good efficacy of COVID-19 vaccines.

**Methods:** A prospective, comparative cohort study was conducted in Beijing Ditan Hospital to evaluate the clinical, immunological and genomic characteristics of COVID-19 breakthrough infections. Data on 88 COVID-19 breakthrough cases (vaccinated group) and 41 unvaccinated cases (unvaccinated group) from June 1 to August 20, 2021 were extracted from a cloud database. Among these 129 COVID-19 cases, we successfully sequenced 33 whole genomes, including 16 from the vaccinated group and 17 from the unvaccinated group.

**Results:** Asymptomatic and mild cases predominated in both groups, but 2 patients developed severe disease in the unvaccinated group. Between the two groups, the median time of viral shedding in the vaccinated group were significantly lower than those in the unvaccinated group (p = 0.003). A comparison of dynamic IgG titres of cases in the two groups indicated that IgG titres in the vaccinated group showed a significantly increasing trend (P =0.028). The CD4^+^T lymphocyte count was lower in the unvaccinated group, and there was a significant difference between the two groups (p=0.018). In the vaccinated group, the number of moderate cases who received Sinopharm BBIBP (42 cases) was significantly higher than those who received Sinovac Coronavac (p=0.020). Whole-genome sequencing revealed 23 cases of delta variants, including 15 patients from the vaccinated group. However, no significant difference was observed in either the RT-qPCR results or viral shedding time.

**Conclusions:** COVID-19 vaccine breakthrough infections were mainly asymptomatic and mild, the IgG titres were significantly higher and increased rapidly, and the viral shedding was short. Delta variants may be more likely to cause breakthrough infections, and vaccination may not reduce the viral loads and shedding time.

## Introduction

Coronavirus disease 2019 (COVID-19) has now become a significant public health concern worldwide. Globally, as of 5:08pm CET, 17 November 2021, there have been 254,256,432 confirmed cases of COVID-19, including 5,112,461 deaths, reported to WHO [1]. Through prevention, control, and treatment, the pandemic situation in China is under control, but there are still local outbreaks and imported cases in some areas.

The mainly vaccines in China are inactivated vaccine, adenovirus vector vaccine and recombinant subunit vaccine, of which inactivated vaccine covers the most widely. At least 13 different vaccines (across four platforms) have been administered in China. The first mass vaccination program started in early December 2020. As of 14 November 2021, a total of 7,307,892,664 vaccine doses have been administered [1]. COVID-19 vaccines effectively prevent infection, serious illness, and death; however, a fraction of fully vaccinated people still get SARS-CoV-2 infection. An infection that occurs in a fully vaccinated person refers to as a “breakthrough infection” [2]. In the context of widespread vaccine coverage, breakthrough infection has become a new challenge in fighting against COVID-19.

The delta variant is currently the predominant variant of the virus in the world. People infected with the delta variant, including fully vaccinated people with symptomatic breakthrough infections, can transmit the virus to others [3]. If delta variants are more likely to cause breakthrough infections, vaccination may reduce the viral loads, and shedding time has become the main topic recently. At present, there are reports of cases of breakthrough infections worldwide, and in China, there were microscopic researches for vaccine breakthrough infections. In the present study, we summarized the clinical data of the patients with breakthrough infections in Beijing Ditan Hospital, compared them with the unvaccinated group, and further analyses the viral genome, clinical and immunological features.

## Materials and Methods

### Ethical approval

The study was approved by the Institutional Review Board of Beijing Ditan Hospital, Capital Medical University in Beijing (approval number JDLY2020-020-01).

### Study design and recruitment of cases

Since January 2020, we established the cloud database of COVID-19 patients in Beijing Ditan Hospital, as of 5 October 2021, 802 confirmed cases and 175 asymptomatic infections were prospectively analysed. In this study we extracted a total of all 129 adult COVID-19 cases from June to August 2021, and the cases were divided into the breakthrough infection group which were vaccinated with different types of vaccines and the unvaccinated group. COVID-19 patients were diagnosed based on the 8th version of the Diagnosis and Treatment Protocol for Novel Coronavirus Pneumonia Patients[4], and the clinical severity was categorized into 4 grades: mild (the clinical symptoms were mild, and there was no evidence of pneumonia on imaging), moderate (fever and respiratory symptoms, imaging manifestations of pneumonia), severe (dyspnoea with a respiratory rate of 30/min, hypoxemia with oxygen saturation of 93%, PaO2/FiO2 of 300 mm Hg, if the clinical symptoms had gradually worsened, and the lung imaging showed that the lesions progressed more than 50% within 24 to 48 h), and critical (developed complications, including respiratory failure that needed mechanical ventilation, shock, or other organ failure that needed intensive care unit [ICU] monitoring and treatment).

Asymptomatic patients with no clinical symptoms but positive RT-qPCR results were also admitted to the hospital. As of this study, all of the patients have been discharged.

### Data collection

The data of the 129 COVID-19 cases from the two groups were extracted from the cloud database. Medical record reviews of the patients were performed to collect the patients’ underlying medical conditions, laboratory test results, and hospital course. In addition, epidemiological data of the patients were collected. The antibodies levels of all patients were followed up to the 8th week after diagnosis.

### Laboratory testing

Oropharyngeal swabs, nasopharyngeal swabs, or sputum specimens obtained from the patients during their hospital stay were collected for RT-qPCR testing. RT-qPCR was conducted using the SARS-CoV-2 nucleic acid test kit (BioGerm, Shanghai, China) with a fluorescence PCR detector following the manufacturers’ instructions, and the results were showed as the cycle threshold (Ct) for the ORF1ab and N genes of SARS-CoV-2. Corresponding serum samples were tested for anti-SARS-CoV-2 antibodies using a chemiluminescence immunoassay (CLIA; Bioscience, Chongqing, China).

### Whole-genome sequencing and analysis

The selected swab samples of the patients were then sent to the Beijing CDC for further whole-genome sequencing. The viral RNA was extracted using Kingfisher Flex Purification System (ThermoFisher, Waltham, MA, USA), and Libraries were prepared using a Nextera XT Library Prep Kit (Illumina, San Diego, CA, USA), and the resulting DNA libraries were sequenced on a MiniSeq platform (Illumina) using a 300-cycle reagent kit. Mapped assemblies were generated using the SARS-CoV-2 genome (accession number NC_045512) as a reference. Variant calling, genome alignment, and sequence illustrations were generated with CLCBio software. Whole-genome sequence alignment was conducted using the Muscle tool in MEGA (v7.0). A neighbour-joining phylogenetic tree was constructed using MEGA (v7.0), and the Kimura 2-parameter model with 1,000 bootstrap replicates was used. Genomic lineage designation was performed using the “PANGO lineage” typing method (https://cov-lineages.org/).

### Statistical analysis

The statistical analyses were performed using SPSS version 25.0 (SPSS IBM, Armonk, NY, USA). From each individual patient, we entered the demographic and clinical variables. Normal continuous variables are represented by the mean and standard deviation. Student’s t test was used to test for significant differences. Nonnormally distributed continuous variables are represented by the medians and interquartile ranges, and the Mann-Whitney U test was used for comparisons. Categorical variables are expressed as numbers and percentages, and the chi-squared test and Fisher’s exact test were used to compare them. All tests were two-tailed, and statistical significance was defined as a P value lower than 0.05.

## Results

### Basic information of the COVID-19 cases

Of the 129 cases, 8 were indigenous cases with one chain of transmission from China, and the remaining 121 were imported cases from outside China (see Table 1). There were 88 cases of breakthrough infection and 41 control cases. There were 61 males and 27 females in the vaccinated group and 28 males and 13 females in the unvaccinated group (see Table 2).

**Table 1.** Countries of variant origin in the two groups.

| Country of origin | Breakthrough | Unvaccinated |
| --- | --- | --- |
|  | infections | group |
|  | (n=88) | (n=41) |
| China | 6 | 2 |
| Russia | 1 | 3 |
| United States of<br>America | 4 | 0 |
| United Kingdom | 6 | 5 |
| Sweden | 2 | 2 |
| Denmark | 2 | 1 |
| Italy | 0 | 1 |
| Greece | 0 | 6 |
| Hungary | 1 | 0 |
| Serbia | 4 | 0 |
| United Arab<br>Emirates | 23 | 12 |
| Philippines | 28 | 9 |
| Lebanon | 1 | 0 |
| Turkmenistan | 3 | 0 |
| Mongolia | 2 | 0 |
| Sudan | 1 | 0 |
| Senegal | 1 | 0 |
| Zimbabwe | 2 | 0 |
| Papa New Guinea | 1 | 0 |

**Table 2.**
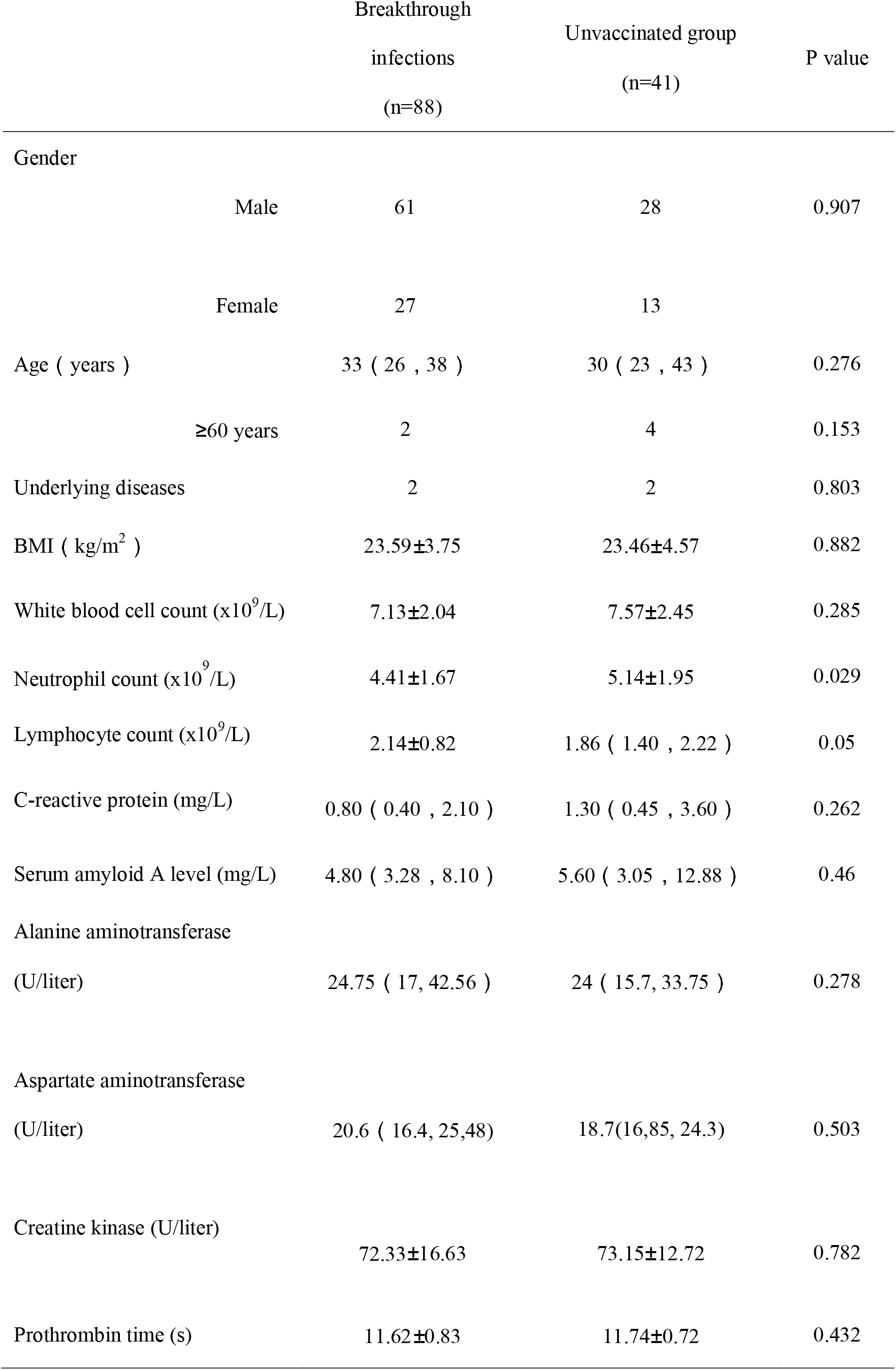
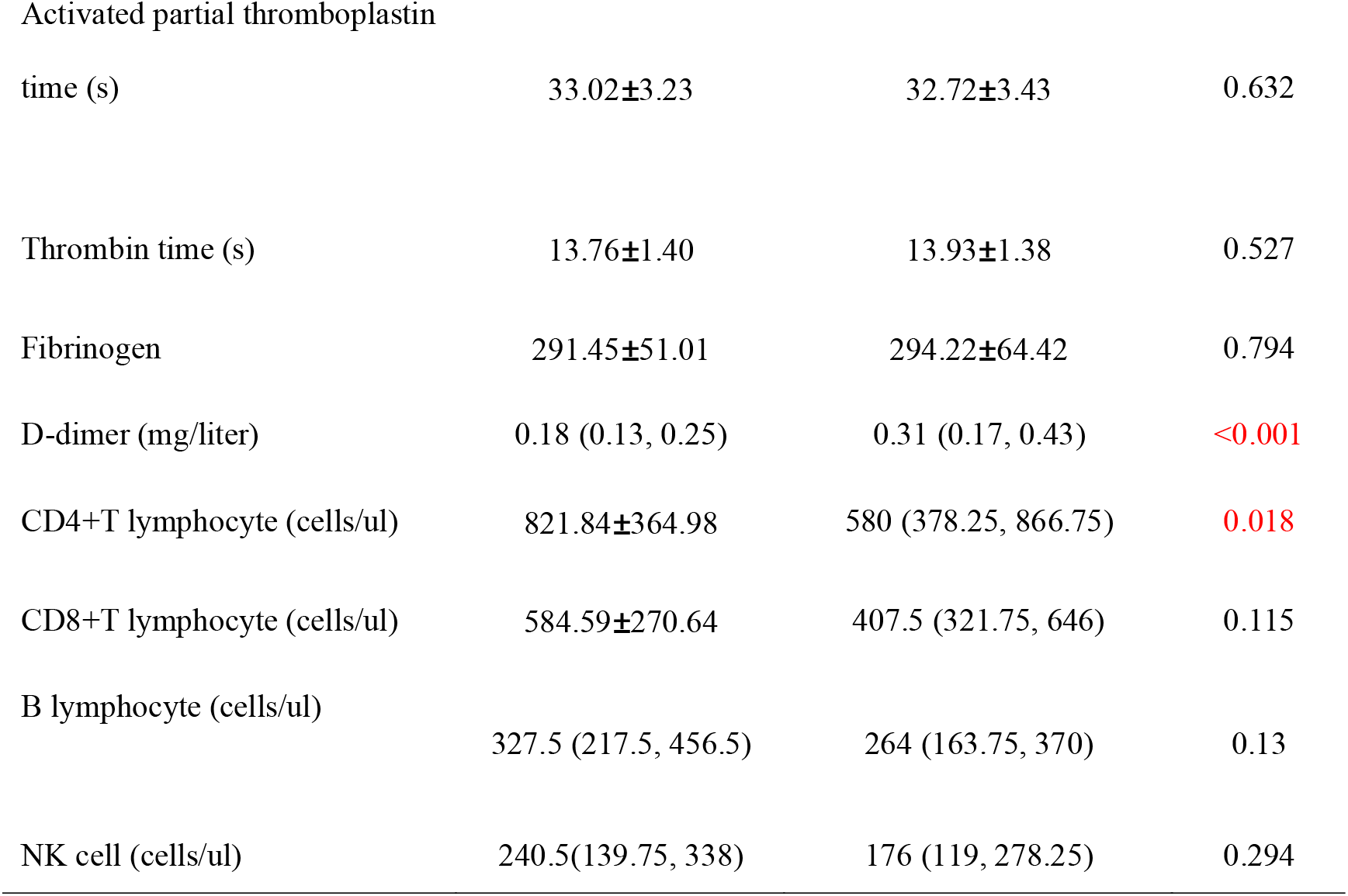
General information and basic laboratory tests in the two groups.

### Whole-genome sequencing and analysis

Clinical samples from all 129 COVID-19 cases were acquired for further sequencing. Since the viral loads were relatively low in tested samples, a total of 33 whole-genome sequences were obtained that had a genome coverage over 98%, and these samples included 16 from the vaccinated group and 17 from the unvaccinated group. After confirming the sequences of the samples by the Pangolin COVID-19 Lineage Assigner Web application [5, 6] (https://pangolin.cog-uk.io/), the 33 strains contained four kinds of variants, including three kinds of VOCs (alpha, beta and delta variants) and the lineage AZ.2 (sublineage of lineage B.1.1.318, which is currently designated as “alert for further monitoring” by the WHO). Among the 16 SARS-CoV-2 strains from the vaccinated group, all strains except one were delta variants, and one sample contained the alpha variant. There were 2 alpha, 2 beta, and 8 delta variants in the unvaccinated group, and there were also 5 strains from the lineage AZ.2. The phylogenetic tree showed that most of the strains did not cluster together within the same lineage, which shows that the strains were of different countries of origin (Figure 1). The indigenous cases (indicated by a yellow arrow) clustered together because they were related cases. Furthermore, the strains from AZ.2 were all imported from Greece, and they formed a clade with a high bootstrap value.

**Figure 1.**
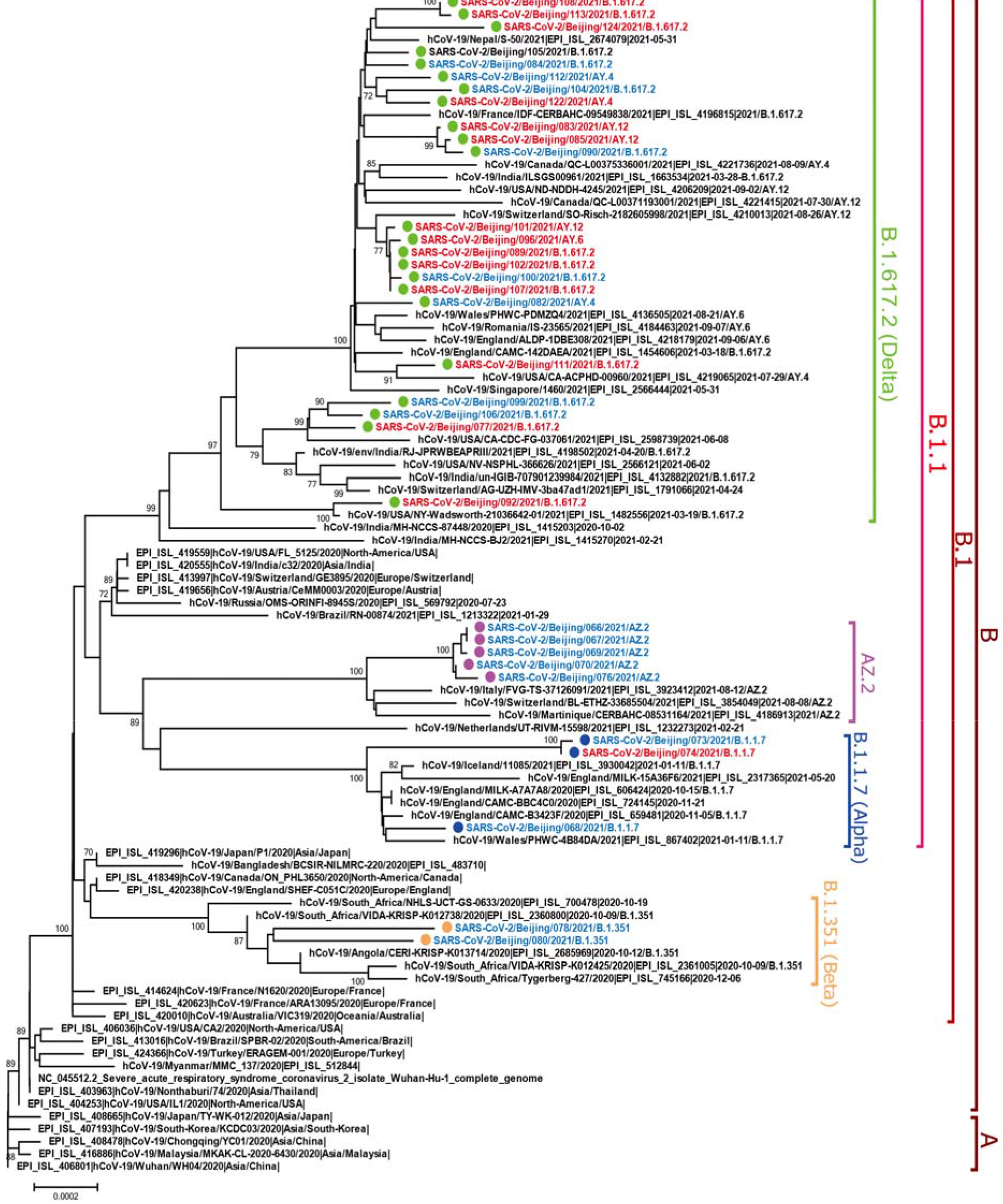
Neighbor-Joining phylogenetic tree based on the whole genome sequences of the SARS-CoV-2. The strains from the vaccine breakthrough infections are indicated with red font; while the strains from unvaccinated infections are indicated with blue font. The PANGO lineages were marked and colored on the right. The tree was rooted using strain WH04 (EPI_ISL_406801) in accord with the root of PANGO tree.

The numbers of nucleotide mutations (including substitutions, insertions and deletions) of the alpha, beta, delta and AY.2 variants ranged from 53 to 66, 50 to 53, 42 to 63 and 67 to 73, respectively. The numbers of amino acid mutations within the spike protein ranged from 11 to 12, 12 to 13, 8 to 15 and 7 to 8, which is suggestive of a high rate of mutation.

### Comparison of the general information and basic laboratory tests in the two groups

The median age of the vaccinated group was 33 (26, 38) years, the median age of the unvaccinated group was 30 (23, 43) years, and there was no significant difference between the two groups (P = 0.276). Two of the patients from each group had underlying diseases. The average body mass index (BMI) of the vaccinated group was 23.59 ± 3.75 kg/m^2^, and that of the unvaccinated group was 23.46 ± 4.57 kg/m^2^. There was no significant difference between the two groups. There were no differences in sex, age, underlying diseases, or BMI between the two groups (Table 1).

There were no significant differences in the laboratory tests between the two groups. These laboratory values included the leukocyte count, lymphocyte count, C-reactive protein (CRP) level, serum amyloid A (SAA) level, Alanine aminotransferase (ALT), Aspartate aminotransferase (AST), Creatine kinase (Cr), Prothrombin time (PT), Activated partial thromboplastin time (APTT), Thrombin time (TT) and Fibrinogen (see Table 1). In addition, the average neutrophil count and D-dimer in the unvaccinated group were higher than that in the vaccinated group, but they were all within the normal range.

The CD4^+^T lymphocyte count, CD8^+^ T lymphocyte count, B lymphocyte count, and NK cell were all lower in the unvaccinated group. The CD4^+^T lymphocyte count was 580 (378.25, 866.75), which was much lower than the breakthrough infections group (821.84±364.98 cells/ul), and there was a significant difference between the two groups (p=0.018) (Table 2).

### Comparison of the clinical severity level in the two groups

The two groups’ patients were mainly either asymptomatic or mild cases, there were 64 asymptomatic patients and 7 mild patients in the vaccinated group, and there were 25 asymptomatic patients and 5 mild patients in the unvaccinated group. There was no significant difference between the two groups (P = 0.335). There were 17 and 9 moderate patients, respectively, and there was no significant difference between the groups (P = 0.729). Two patients in the unvaccinated group developed the severe disease (severe or critical), while no severe patients were in the vaccinated group (Table 3).

**Table 3.** Comparisons of the clinical severity levels in the two groups.

|  | Breakthrough<br>infections<br>( n=88 ) | Unvaccinated<br>group<br>( n=41 ) | P value |
| --- | --- | --- | --- |
| Asymptomatic patients | 64 | 25 | 0.179 |
| Mild patients | 7 | 5 | 0.655 |
| Moderate patients | 17 | 9 | 0.729 |
| Severe or critical patients | 0 | 2 | 0.186 |

### Comparison of viral shedding time, Ct value, IgM and IgG in the two groups

The median time from complete vaccination to the first positive nucleic acid test was 100.50 (24.00, 171.50) days. The median time of viral shedding was 3.00 (1.00, 11.75) days in the vaccinated group and 11.00 (2.00, 23.50) days in the unvaccinated group. There was a significant difference between the two groups (P = 0.003). The viral shedding time was significantly shorter than that of the unvaccinated group. At admission, there were no significant differences between the two groups regarding the median Ct values of the ORF1abgene and N gene (P = 0.148) (Table 4).

**Table 4.** Comparison of the viral shedding time, Ct valuein the two groups at admission.

|  | Breakthrough infections<br>( n=88 ) | Unvaccinated group<br>( n=41 ) | P value |
| --- | --- | --- | --- |
| Time from full vaccination to first | 100.50 ( 24.00 , 171.5 ) |  |  |
| positive nucleic acid test (days) |  |  |  |
| Viral shedding time (days) | 3.00 ( 1.00 , 11.75 ) | 11.00 ( 2.00 , 23.50 ) | 0.003 |
| Ct value of the ORF1ab gene | 35.75 ( 31.33 , 37.77 ) | 34.29 ( 25.46 , 38.03 ) | 0.267 |
| Ct value of the N gene | 35.40 ( 32.80 , 37.40 ) | 34.40 ( 24.20 , 37.40 ) | 0.148 |
| IgM titre | 0.52 ( 0.14 , 1.12 ) | 0.25 ( 0.05 , 0.86 ) | 0.037 |
| IgG titre | 21.85 ( 9.56 , 65.35 ) | 2.20 ( 0.04 , 8.74 ) | 0.000 |

There was no significant difference in IgM titres between the two groups (P = 0.037). The IgG titre ranged from 0.45 to 393.24 in the vaccinated group, with a median of 21.85 (9.56, 65.35), significantly higher than in the unvaccinated group. There was a significant difference between the two groups (P = 0.000) (Table 4). The IgG titres of the 2 severe patients in the unvaccinated group were negative at admission.

### Dynamical comparison of the Ct value, IgM and IgG of COVID-19 patients in the two groups

The IgM titres in the breakthrough infections declined after reaching peaks during the second week after admission, and the IgM titres increased till the sixth week in the unvaccinated group. The IgM titres were higher in the unvaccinated group, and there was a significant difference between the two groups (P = 0.012). The IgG titres in the breakthrough infections gradually increased after admission, and after the fourth week the titres were sharply rose. The IgG titres in breakthrough infections were higher than unvaccinated group at admission and during the whole follow up period, and there was a significant difference between the two groups (P = 0.004) (see Fig.2).

**Figure 2.**
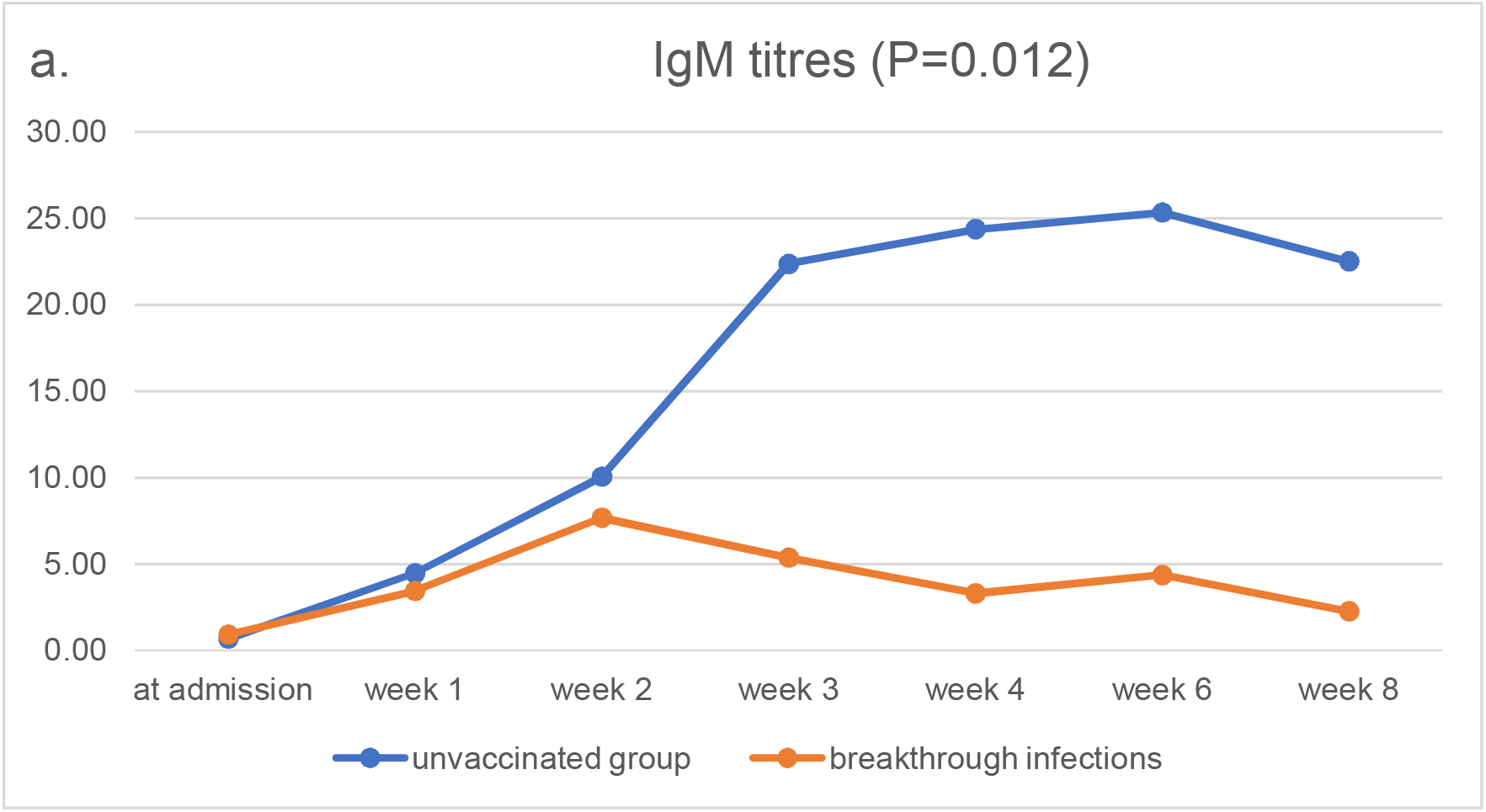
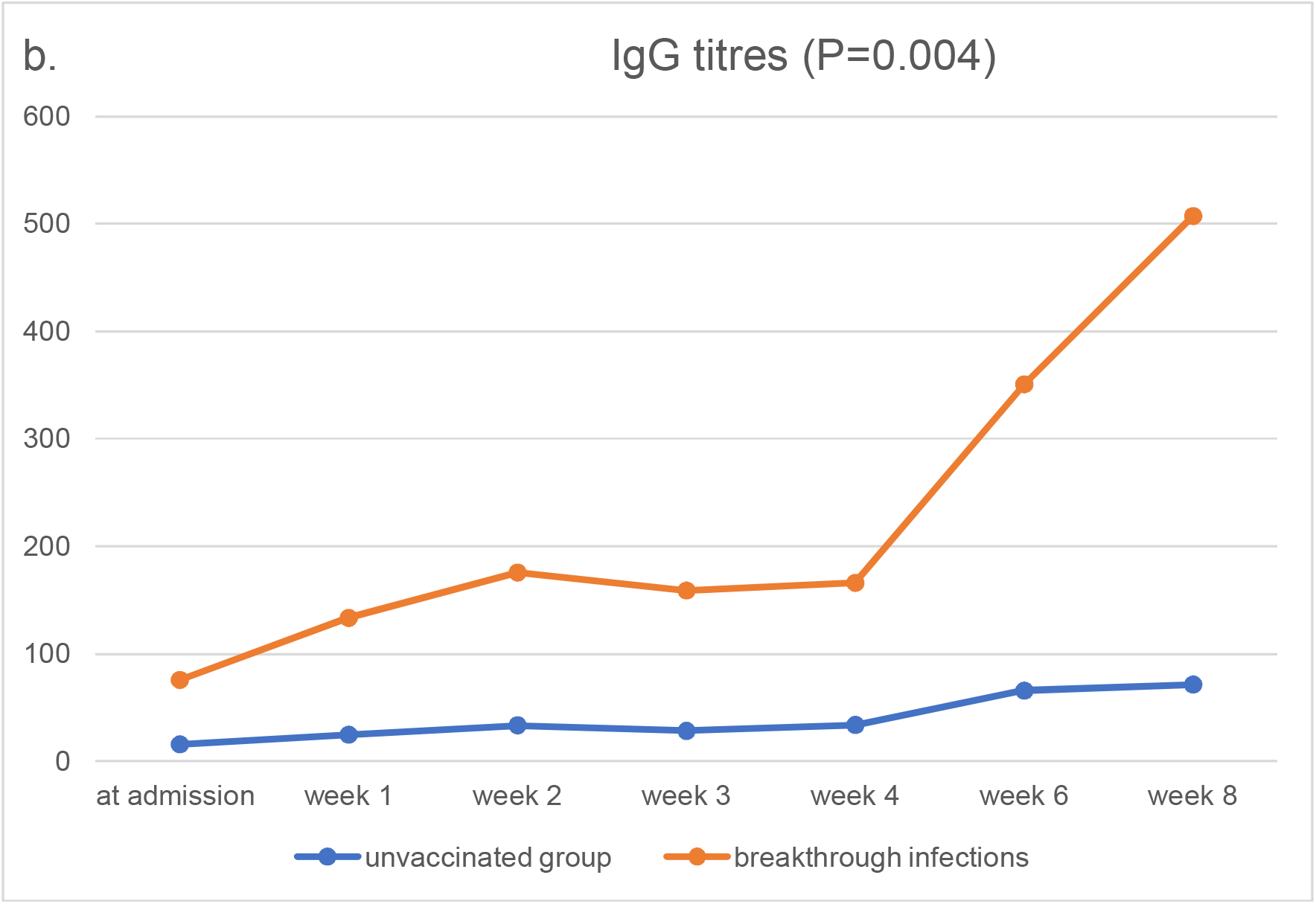
Dynamic comparison of IgM and IgG titres in COVID-19 patients with breakthrough infections and unvaccinated group. a.The IgM titres in the breakthrough infections declined after reaching peaks during the second week after admission, and the IgM titres increased till the sixth week in the unvaccinated group. The IgM titres were higher in the unvaccinated group, and there was a significant difference between the two groups (P = 0.012). b.The IgG titres in the breakthrough infections gradually increased after admission, and after the fourth week the titres were sharply rose. The IgG titres in breakthrough infections were higher than unvaccinated group, and there was a significant difference between the two groups (P = 0.004).

The Ct values of the ORF1ab gene and the N gene at admission and during the second week, third week, and fourth week after admission gradually increased until negative results in the two groups. There were no significant differences between the two groups (P=0.740 and 0.696) (Figure 3).

**Figure 3.**
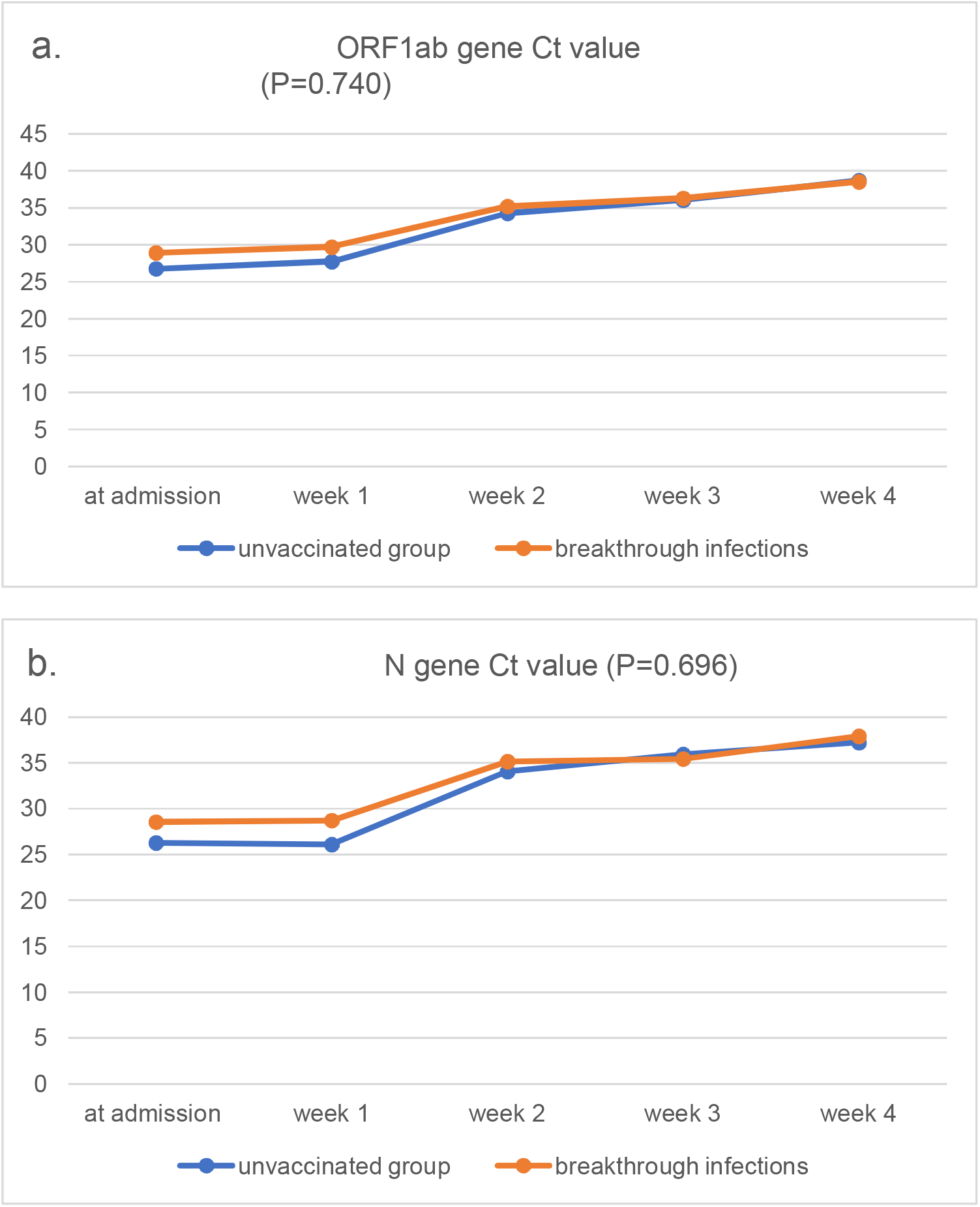
Dynamic comparison of the Ct value of the ORF1ab and N gene in patients with breakthrough infections and unvaccinated group. a.The Ct values of the ORF1ab gene at admission and during the second week, third week, and fourth week after admission gradually increased, and there was no significant differences between the two groups (P=0.740). b.The Ct values of the N gene at admission and during the second week, third week, and fourth week after admission gradually increased, and there was no significant differences between the two groups (P=0.696).

### Comparison of vaccines from different platforms

Among the 88 patients with breakthrough infections, 42 were vaccinated with the Sinopharm BBIBP vaccine, 32 were vaccinated with the Sinovac CoronaVac vaccine, 4 were vaccinated with the Pfizer vaccine (BNT162B2), 1 was vaccinated with the AstraZeneca vaccine, 1 was vaccinated with the Sputnik V vaccine, and in 8 patients, the type of vaccine was unknown. We compared two Chinese inactivated vaccines. There were fewer asymptomatic infections and more moderate patients who had the Sinopharm BBIBP vaccine than patients vaccinated with the Sinovac CoronaVac vaccine, and there was a significant difference between the two groups. However, the Ct value, serum antibody titre, and viral shedding time were similar between the groups (see Table 5).

**Table 5.** Comparison of the Sinopharm BBIBP vaccine and the Sinovac CoronaVac vaccine.

|  | Sinopharm BBIBP<br>(n=42) | Sinovac CoronaVac<br>(n=32) | P value |
| --- | --- | --- | --- |
| Asymptomatic patients | 25 | 27 | 0.020 |
| Mild patients | 4 | 3 | 1.000 |
| Moderate patients | 13 | 2 | 0.020 |
| Ct value of the ORF1ab gene | 35.80 ( 32 , 37.22 ) | 36.02 ( 27.89 , 38.86 ) | 0.495 |
| Ct value of the N gene | 34.70 ( 31.99 , 37.3 ) | 36.33 ( 28.1 , 37.43 ) | 0.543 |
| IgM titre | 0.67 ( 0.13 , 1.65 ) | 0.45 ( 0.16 , 1.34 ) | 0.513 |
| IgG titre | 32.04 ( 5.73 , 108.28 ) | 16.19 ( 9.14 , 22.92 ) | 0.111 |
| Viral shedding time (days) | 3.00 ( 1.00 , 11.50 ) | 3.00 ( 1.00 , 12.00 ) | 0.699 |
| Time from full vaccination to first positive nucleic acid test (days) | 176.79±95.53 | 23.50 ( 15.50 , 75.25 ) | <0.001 |

### Comparison of the breakthrough infections and unvaccinated group in the patients with the delta variant

Among the patients with the delta variant, there were 15 breakthrough infections and 8 patients were unvaccinated. There were no differences in the Ct values of the ORF1ab and N genes between the two groups (P = 0.857 and 0.794), and there was no significant difference in the viral shedding time between the two groups (P = 0.352). The IgM and IgG titres in patients with breakthrough infections were higher than those without breakthrough infections (P=0.007 and P=0.001) (see Table 6).

**Table 6.** Comparison of patients with breakthrough infections and controls with the delta variant

|  | Breakthrough<br>infections with the<br>delta variant (n=15) | Controls with the<br>delta variant (n=8) | P value |
| --- | --- | --- | --- |
| Asymptomatic<br>patients | 3 | 2 | 1.000 |
| Mild patients | 4 | 2 | 1.000 |
| Moderate patients | 8 | 4 | 1.000 |
| Ct value of the<br>ORF1ab gene | 26.52±6.52 | 26.05±4.13 | 0.857 |
| Ct value of the N gene | 25.18±6.41 | 24.44±6.20 | 0.794 |
| IgM titre | 0.22 (0.08, 0.65) | 0.05 (0.03, 0.10) | 0.007 |
| IgG titre | 4.87 (0.93, 38.53) | 0.02 (0.02, 0.05) | 0.001 |
| Viral shedding time<br>(days) | 22.11±5.73 | 25.5 (25, 26) | 0.352 |

## Discussion

### Breakthrough infections

Some evidence suggests that patients who have recovered from COVID-19 can be reinfected [7-9]; Thus, vaccination may not lead to long-term and effective immunity against SARS-CoV-2 [8].

Pediatric patients with COVID-19 may experience milder illness with atypical clinical manifestations. These patients rarely have lymphopenia [10], so we excluded them in this study. We characterized the patients with SARS-CoV-2 breakthrough infections among 88 fully vaccinated patients, six of them had domestically confirmed SARS-CoV-2 strains from China, and the other strains imported from aboard. Most of the patients were either asymptomatic or had a mild case of COVID-19; no patients progressed to severe disease. The Ct values of the ORF1ab gene and the N gene between the patients with breakthrough infections and the unvaccinated group were similar, but the average duration of viral shedding in patients with breakthrough infections was much lower than that in the unvaccinated group. These data indicate that COVID-19 vaccines are still effective at preventing severe illness and death.

Similar cases have occurred in other countries. During July 2021, 469 cases of COVID-19 were identified among Massachusetts residents; the vaccination coverage among eligible Massachusetts residents was 69%. Approximately three-quarters (346; 74%) of cases occurred in fully vaccinated persons (those patients who had completed a 2-dose course of an mRNA vaccine). The Ct values were similar among the samples from the fully vaccinated patients and the samples from those who were not fully vaccinated [11]. According to Bergwerk et al., 39 breakthrough cases were identified among fully vaccinated health care workers. They concluded that the occurrence of breakthrough infections with SARS-CoV-2 correlated with neutralizing antibody titres during the peri-infection period, and most breakthrough infections were mild or asymptomatic cases [12]. Guangzhou, China, reported 38 imported COVID-19 patients who had received inactivated vaccines (Vero cells). Patients infected with SARS-CoV-2 after a vaccination can produce IgG antibodies rapidly in the early stage of disease compared to those who did not receive an inactivated vaccine, and RT-qPCR tests stay negative for a longer period of time [13].

### Immunological characteristics

In this study, the median time from fully vaccinated to positive RT-qPCR was 100.5 days, and the median IgG titre of patients with breakthrough infections was 21.85 at admission, indicating that the antibodies can still be detected approximately three months after vaccination, and the decrease in the antibody titre may be related to the development of breakthrough infections. The reduction in antibody titre may weaken the protective effect against infection and infectivity. The IgG levels in the vaccination group gradually increased after admission and were significantly higher than that in the unvaccinated group at admission, during the second and the third weeks.

The modelling of the data regarding the neutralizing antibody titres and vaccine effectiveness suggested that the neutralizing antibody titre needed to prevent severe illness is lower than the titre required to avoid infection. Although we do not yet know the target titre to prevent infection, it is estimated to be higher than the titre needed to prevent severe illness [14]. Khoury et al. assumed that after eight months following infection, the decay rate would slow down. They modelled the decay rate of the neutralization titre, assuming that it slowed linearly with a 10-year half-life over 1, 1.5, or 2 years (details are presented in their Methods section). Their analysis predicted that even without vaccine boosters, a significant proportion of individuals might maintain a long-term protection from severe infection by an antigenically similar strain, even though these patients may become susceptible to mild disease [15].

Vaccine-elicited neutralizing antibody titres correlated with the protective efficacy, suggesting that there was immunity and protection, and the presence of neutralizing antibodies from a prior infection was significantly associated with the protection against reinfection [16,17].

Researchers have found a positive correlation between the neutralizing antibody titres and the levels of IgG antibodies. The levels of neutralizing antibodies gradually decrease over for 3 months. Therefore, there is an obvious risk for the possibility of reinfection that is related to the fast decay of antibodies or secondary to a poor elicitation of neutralizing antibodies [18,19].

Moreover, our study found that the vaccinated group had higher counts of CD4^+^T lymphocytes, CD8^+^T lymphocytes, B lymphocytes, and NK cells. T-cell-mediated adaptive immune responses are essential for viral clearance and long-term antiviral immunity but may contribute to cytokine storm and could compromise in severe cases of COVID-19 patients due to T-cell exhaustion [20]. Successful vaccines should generate SARS-CoV-2-reactive T cells with high specificity for potent immune responses devoid of the undesired effects of inflammation or inception of disease. There are many ongoing vaccine trials, but it is unknown whether they will provide long-lasting protective immunity. Most vaccines are designed to induce antibodies to the SARS-CoV-2 spike protein, but it is unknown if this will be sufficient to induce complete protective immunity to SARS-CoV-2 [21].

### Delta variant

In this study, there were 9 breakthrough infections, and 7 unvaccinated patients had the delta variant. Compared with unvaccinated patients, there was no difference in the Ct values between the two groups, and there was no significant difference in viral shedding time between the two groups.

The B.1.617.2 (delta) variant was first detected in India in December 2020 and became the most commonly reported variant starting in mid-April 2021 [22]. Mutations of the delta variant may have higher replication rates, which leads to higher viral loads and increased transmission rates [23]. Recently, one study reported only a slight difference in the vaccine efficacy against delta variant and alpha variant after patients received two doses of the vaccine [24].

This study had some limitations. There were relatively few cases in the unvaccinated group, and most patients did not have full genome sequencing results, which may be related to the low viral load (high CT value) in some patients which were tested negative in the origin country. In addition, the number of patients that had breakthrough infections after being vaccinated with non-inactive vaccines was much lower than the number of patients vaccinated with inactive vaccines.

Being vaccinated does not mean we can throw caution to the wind and put ourselves and others at risk, mainly because the research is still ongoing into how much vaccines protect against disease, infection, and viral transmission. Therefore, for the foreseeable future, we still need to continue wearing masks and washing our hands, and we need to ensure good indoor ventilation, continue physical distancing, and continue avoiding crowds.

## Data Availability

All data produced in the present study are available upon reasonable request to the authors
All data produced in the present work are contained in the manuscript

## ACKNOWLEDGMENTS

We thank all physicians who participated in the management of these COVID-19 cases. This study was funded by Chang Jiang Scholars program (grant no.2019077). W.X, Z.C. conceived the study, W.X., Z.C. designed the study, Z.C. and W.X. and P.Y. maintained the database for data collection, Z.C., W.X. supervised the data collection, D.T., Y.S., M.Z., Y.P., Z.G., Y.Z., X.R., J.W. and Y.X. interpreted the data, D.T., Y.S. and M.Z. did the statistical analysis, D.T. prepared the manuscript, D.T., Y.S., M.Z. and Z.G. wrote the manuscript, D.T. and Z.G. prepared the figures, M.Z. and Y.P. and P.Y. did the COVID-19 specimen processing and sequencing, and all authors reviewed and approved the final version of the manuscript. Z.C., W.X., and P.Y. are the guarantors.

We declare no conflicts of interest.

## Notes

### Competing Interest Statement

The authors have declared no competing interest.

### Author Declarations

Institutional Review Board of Beijing Ditan Hospital, Capital Medical University in Beijing gave ethical approval for this work (approval number JDLY2020-020-01).

